# The Introduction of Social Health Insurance and Health Care Seeking Behavior in Urban Ethiopia

**DOI:** 10.1101/2023.07.05.23292262

**Authors:** Zahra Zarepour, Anagaw Mebratie, Dessalegn Shamebo, Zemzem Shigute, Getnet Alemu, Arjun S. Bedi

## Abstract

**Objectives:** In recent years, to enhance access to and use of health care the government of Ethiopia has introduced voluntary Community Based Health Insurance (CBHI) schemes for the rural and informal sectors of the economy. After years of planning and the ratification of a legal framework the government proposes to introduce a compulsory Social Health Insurance (SHI) program for formal sector employees. The proposed scheme will provide access to contracted health care facilities at a premium of 3% of the gross monthly income of employees with another 3% coming from the employer. While several studies have examined the willingness to pay this premium, little is known about the health care seeking behaviour (HSB) of formal sector employees. In part, the implementation of the SHI has been delayed due to the unwillingness of public servants to pay the proposed premium. Scheme coverage which will be restricted to contracted facilities, may also be contentious if this is dominated by publicly provided health care services. This paper investigates both, the determinants of health care seeking behaviour of formal sector employees and their families and attitudes related to the introduction of SHI such as fairness, affordability, and willingness to pay the SHI premium. Through these explorations, the paper sheds light on the potential challenges for the implementation of SHI.

**Setting:** The study is based on a survey of formal sector employees and their families in urban Ethiopia. It covers the major administrative regions of the country and contains information on 2,749 formal sector employees and their families or a total of 6,894 individuals.

**Results:** Regarding outpatient care, conditional on falling ill, 85.5% sought some form of care within a couple of days (2.4 days) of falling ill. The bulk (94%) of those who did seek care, opted for formal treatment. A majority of the visits (55.9%) were to private health clinics or hospitals. In the case of inpatient care, the picture was reversed with a majority of health care seekers visiting public sector hospitals (62.5%). There is a strong positive link between income and the use of private health services. A majority of the sample (67%) supported the introduction of SHI but only about 24% were willing to pay a premium of 3% of their gross monthly income. The average WTP was 1.6%. Respondents in the two richest income quintiles were far more likely to oppose SHI and consider it unfair.

**Conclusion:** The prominent role of the private sector especially in terms of outpatient care and the stronger resistance to SHI amongst the two richest income quintiles, that is, those who are most likely to use private health care providers, suggests that the SHI program needs to actively include private health care facilities within its ambit. Additionally, as was done prior to the introduction of the CBHI, concerted efforts at enhancing the quality of care available at public health facilities, both, in terms of perception and patient-centred care and in terms of addressing drug and equipment availability bottlenecks, are needed. A combination of these two measures is likely to enhance support for the introduction of SHI.

## Introduction

In the past 20 years, Ethiopia has witnessed a sharp expansion in its public healthcare system. Between 2000 and 2021 there has been a 22-fold increase in the number of health posts, an 11-fold increase in the number of health centres and a four-fold increase in the number of public hospitals [1-2]. Per capita healthcare spending has grown from US$5.4 in 2000 to US$36.3 in 2020 (including COVID-19 spending) [3]. Due to such efforts as well as changes in healthcare seeking behaviour, access to essential healthcare services, as measured by the availability of health facilities within a two-hour walking distance has increased from 50.7% in 2000 to more than 90% in 2019 and outpatient (OPD) attendance per capita has increased from 0.27 in 2000 to 1.02 visits in 2020 [4-5].

At the same time as these supply-side investments, the country has made remarkable progress on the demand-side through the implementation of voluntary Community-based Health Insurance (CBHI) schemes for the rural and informal sectors of the economy. Based on figures from a presentation made by the Ethiopian Health Insurance Agency (EHIA), since 2013, the government has steadily expanded the scheme and as of June 2022, CBHI schemes have been implemented in 950 rural districts and urban centres covering about 46.5%% of Ethiopia’s 22 million households. More than 70% of the target population is enrolled, which is high, as compared to other voluntary CBHI schemes in Sub-Saharan Africa. The renewal rate is 82% [6]. While far from universal and shy of the target set by the government [7], the CBHI scheme continues to spread and various steps to enhance the sustainability of the scheme are on-going.

In marked contrast, a compulsory Social Health Insurance scheme intended for the formal sector of the economy, which was proclaimed in the federal government’s gazette more than a decade ago [8] is still awaiting introduction. The proposed scheme sought to cover current and former (pensioners) public sector employees including civil servants, employees of public sector enterprises; non-government organisations, religious institutions, and private sector firms with ten or more employees. The scheme envisaged coverage of essential health services and other critical curative services at any health facility that had concluded an agreement with the EHIA - the agency established to administer the scheme.

While scheme launch has often seemed imminent, it has been repeatedly delayed due to two main reasons. First, civil servants and employees of public sector enterprises have expressed their unwillingness to pay the proposed SHI premium of 3% of their gross salaries. While there are no national-level studies, analysis of specific groups of formal sector employees in various parts of the country provides a picture of resistance. For instance, analysis of six focus group discussions in Addis Ababa suggests that in the case of specific benefit packages, participants from public sector enterprises are willing to pay 3% of their monthly salaries while civil servants offer 0.5% [9]. More formal willingness to pay (WTP) studies conducted in Addis Ababa on various samples (ranging from 250 to 503) of civil servants reveal that between 17% to 35% of the sampled respondents are willing to pay the 3% premium. According to these studies, the mean willingness to pay ranges between 1.5% to 2.5% of gross monthly salaries [10-13]. Analysis of data from other cities (Bahir Dar, Gondar, Dessie City, Mujja) reveals a similar picture with willingness to pay the 3% premium ranging from a low of 17.3% of the 488 sampled civil servants in Bahir Dar to a high of 37.6% of the 375 civil servants sampled in Mujja town [14-17]. In contrast to the low expressed WTP in these studies, a handful of papers provide a more sanguine picture. For instance, in Mekelle, the largest city in the country’s Tigray region, a majority (85%) of the 381 public servants were willing to be part of the social health insurance scheme, with a mean WTP of 3.6% of their monthly salary [18]. Although not as high as in Mekelle, surveys amongst teachers in Wolaita Sodo and in Gondar towns revealed WTP rates of 74% and 62%, respectively [19-20].

The second reason for the delay is related to the scheme’s heath care coverage. In principle, the SHI could cover costs in both private and public health facilities, as long as there is a contract in place. However, there is a fear that the scheme may restrict coverage to public health facilities rather than covering potentially higher quality private care [21]. This in turn may lead to crowding of public health care facilities and/or drive up the costs of health care as formal sector employees may have to pay for SHI and still incur out-of-pocket (OOP) expenditure as they seek care in non-contracted private facilities. Unlike the body of literature which has investigated willingness to pay amongst those targeted by the SHI, information on health-seeking behaviour (HSB) for formal sector employees is limited [22]. The existing literature tends to focus on healthcare seeking behavior for children or for specific diseases such as HIV/AIDS and Tuberculosis [23-26]. Information on the HSB of formal sector employees is needed to shed light on the potential challenges that current patterns of HSB may create for the implementation of the SHI.

This paper contributes to the body of work designed to inform the implementation of the SHI in Ethiopia. Such evidence is clearly needed to direct the efforts of the government and the EHIA as it strives to launch the SHI. This paper responds to such concerns and uses data collected from four major Ethiopian cities and formal sector workers including civil servants, employees of public sector enterprises, employees of private sector enterprises, and pensioners to explore both their HSB and attitudes related to the introduction of SHI such as fairness, affordability, and willingness to pay the SHI premium and thereby to inform the efforts of the EHIA.

## Data and Methods

### Data

This study is based on a retrospective cross sectional household survey, conducted in-person in four main cities of the country. The data were collected in June-July 2016. The cities included in the survey were Addis Ababa – the country’s primate city, Bahir Dar, the largest city in the Amhara region, Hawassa, the largest city in the SNNP region, and Mekelle, the largest city in the country’s Tigray region. These cities were purposively selected as they accounted for about 20 percent of the estimated 4.3 million formal sector employees distributed across five categories - civil servants, public sector enterprise employees, private sector workers, NGO workers, pensioners (former civil servant and public sector enterprise workers).

Power calculations (power of 0.8 and significance of 5%) designed to detect the effect of health insurance on the utilization of inpatient care, outpatient care and out-of-pocket (OOP) health expenditure, yielded sample sizes of 3,000 to 4,200 individuals.^1^ Based on budgetary considerations, a sample size of 2,100 households which was expected to yield more than 4,200 individuals was targeted. The distribution of the sample across cities and type of formal sector employees was designed to represent the population of formal sector workers in these four cities. Population level information on the distribution of formal sector workers across different sectors was available from the Ethiopian Statistical Service and the EHIA. This information was used to distribute the sample size across each sector (see Table 1). However, distribution across both cities and sectors was unavailable for all category of workers. We did, however, have access to information on the distribution of civil servants across each city. This was used as the basis for distributing the sample across the four cities (see Table 2).

**Table 1.** Potential SHI members based on a 2013 labour force survey and estimates from Ethiopian Health Insurance Agency

| Sector | Number of employees<br>(Share of total in %) | Sample distribution-<br>*Planned*<br>(Share of total in %) | Sample distribution-<br>*Achieved*<br>(Share in total in %) |
| --- | --- | --- | --- |
| Government worker/civil servant <sup>a</sup> | 1,410,572<br>(32.8) | 690<br>(32.8) | 1280<br>(46.6) |
| Public sector enterprise <sup>b</sup> | 424,039<br>(9.9) | 208<br>(9.9) | 195<br>(7.1) |
| Private sector enterprise <sup>b</sup> | 1,789,963<br>(41.7) | 872<br>(41.5) | 949<br>(34.5) |
| NGO workers <sup>b</sup> | 84,808<br>(1.98) | 42<br>(2.00) | 92<br>(3.4) |
| Civil servant/public sector enterprise pensioners <sup>a</sup> | 590,024<br>(13.75) | 289<br>(13.76) | 233<br>(8.5) |
| *Total* | *4,290,406* | *2,101* | *2,749* |

**Notes:** <sup>a</sup> From EHIA, actual number of employees in May 2016;
<sup>b</sup> Estimated from Central Statistical Office, labour force survey

**Table 2.**
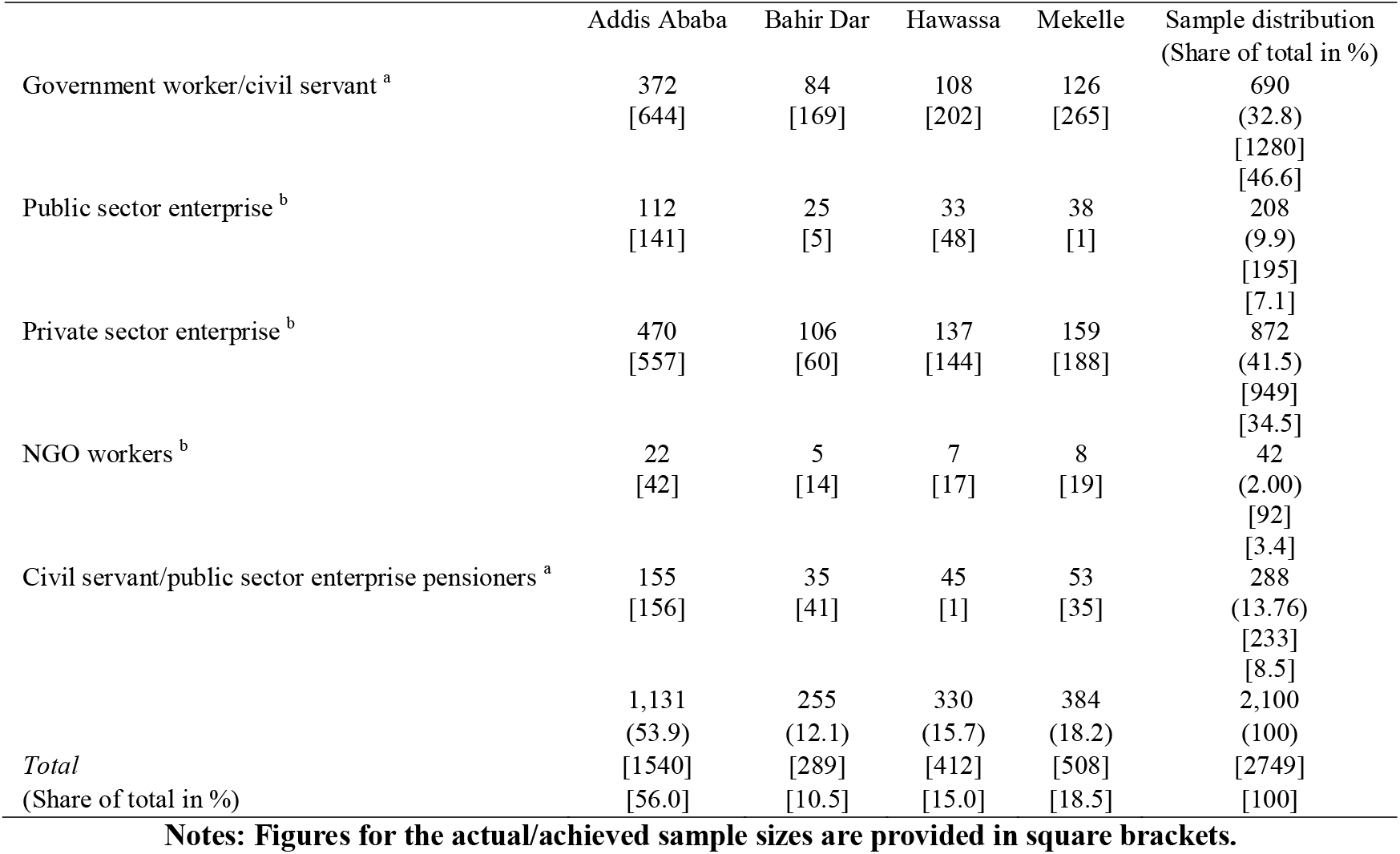
Distribution of sample by sector and city Planned versus Achieved

**Table 3.** Socio-economic characteristics

|  | Full sample | Employees | Public sector | Private/ NGO | Pensioner |
| --- | --- | --- | --- | --- | --- |
| City |  |  |  |  |  |
| Addis Ababa | 0.512 | 0.560 | 0.532 | 0.575 | 0.670 |
| Bahir Dar | 0.115 | 0.105 | 0.118 | 0.071 | 0.176 |
| Hawassa | 0.177 | 0.150 | 0.169 | 0.155 | 0.004 |
| Mekelle | 0.195 | 0.185 | 0.180 | 0.199 | 0.150 |
| Sex (Male=1) | 0.486 | 0.576 | 0.531 | 0.608 | 0.717 |
| Age |  |  |  |  |  |
| Under 18 years old | 0.280 | 0.003 | 0.001 | 0.004 | 0.015 |
| 18-34 years old | 0.399 | 0.505 | 0.464 | 0.619 | 0.524 |
| 35-55 years old | 0.243 | 0.398 | 0.488 | 0.331 | 0.359 |
| More than 55 years old | 0.079 | 0.094 | 0.047 | 0.046 | 0.102 |
| Household size |  |  |  |  |  |
| One-person | 0.069 | 0.152 | 0.143 | 0.178 | 0.094 |
| 2-3 person | 0.243 | 0.314 | 0.292 | 0.332 | 0.369 |
| 4-5 person | 0.454 | 0.385 | 0.410 | 0.346 | 0.395 |
| 6 and more person | 0.234 | 0.150 | 0.155 | 0.144 | 0.142 |
| Monthly income in Birr <sup>a</sup> (household/individual) | 9027.362<br>(8971.435) | 5304.534<br>(5781.062) | 4966.103<br>(5125.078) | 6374.473<br>(6744.358) | 2682.605<br>(3522.130) |
| Education <sup>b</sup> (head of household/individual) |  |  |  |  |  |
| No formal schooling | 0.037 | 0.008 | 0.004 | 0.007 | 0.039 |
| Primary education | 0.154 | 0.063 | 0.039 | 0.054 | 0.266 |
| Secondary education | 0.210 | 0.203 | 0.187 | 0.207 | 0.293 |
| Tertiary or university degree | 0.599 | 0.725 | 0.771 | 0.732 | 0.402 |
| Self-assessment health status |  |  |  |  |  |
| Very poor | 0.001 | 0.001 | 0.000 | 0.001 | 0.004 |
| Poor | 0.011 | 0.014 | 0.010 | 0.010 | 0.060 |
| good | 0.063 | 0.078 | 0.081 | 0.060 | 0.138 |
| Very good | 0.228 | 0.242 | 0.240 | 0.254 | 0.207 |
| Excellent | 0.697 | 0.666 | 0.670 | 0.676 | 0.591 |
| N | 6894 | 2,749 | 1,475 | 1,041 | 233 |

**Note:** Standard deviations are in parentheses; <sup>a</sup> Income refers to the household income for the full sample and to individual incomes in the employee and sector-specific samples. <sup>b</sup> Education refers to education level of the household head in the full sample and to individual education in the employee and sector-specific samples.

Within each city, sample selection was tailored to the sector in question. For instance, with regard to civil servants, in each city, a list of ministries, agencies and bureaus was drawn up and organizations were selected for survey on the basis of probability proportion to their employment size and within each selected organization, respondents were randomly surveyed (see Table A1). A tricky part of the data collection was gathering information on pensioners. In each city the survey team identified the locations where pensioners receive payments and allocated the targeted sample size equally to each of these payment locations. For instance, the Addis Ababa survey team visited the Federal main Post Office which handles payments for pensioners in Addis Ababa. The office provided a list of pensioners and payment centres in ten locations in the city. Five of these locations were randomly selected and an equal number of pensioners were surveyed from each of these locations at the time that they came to receive payments (see Table A2). A similar approach was followed in other cities.

After ensuring that a selected respondent was willing to participate in the survey, enumerators gathered individual and household-level information. The survey contained a household roster which gathered socio-economic information on all household members, their health status and lifestyle choices, outpatient and inpatient health care utilization, financing of heath care, and whether they currently had any form of health insurance. As it turned out, we were able to gather more information and the sample at hand includes 2,749 formal sector employees and their families (6,894 individuals). The key difference (see Table 1) as compared to the plan is a larger share of public sector workers (54% actual versus a plan of 43%) and smaller shares of private sector workers (38% actual versus 43% planned) and pensioners (8% actual versus 14% planned). In terms of the distribution of the sample across the four cities (Addis Ababa - 56% actual versus 54% planned; Bahir Dar - 10.5% planned versus 12.1% actual; Hawassa – 15% actual versus 15.7% planned; Mekelle – 18.5% actual versus 18.2% planned) the differences between the plan and realisation are minor (see Table 2). While not perfect, arguably the sample provides a representative picture of HSB and attitudes towards SHI across formal sector workers residing in these four cities.

### Methods

The paper relies on descriptive statistics, logit and multinomial logit (MNL) models to examine its objectives. Logit models are used to examine the probability of seeking treatment conditional on reporting an illness while multinomial logit models are used to examine choice of health care provider. Logit models are also used to explore attitudes towards SHI. In all instances, the outcomes are treated as functions of socio-demographic traits such as household demographics, education of household head, household income, and the share of a household with health insurance. The models also control for regional fixed effects and in some instances, sector of employment.

### Ethics clearance

Ethics approval (IDPR/LT-0005/2016) was provided by the Research Ethics Committee of the Institute of Development Policy and Research, Addis Ababa University.

## Results

Our discussion of the results begins by commenting on the descriptive statistics of the data, followed by an examination of healthcare seeking behaviour in the case of outpatient care and in-patient care. We end the section by assessing attitudes towards the introduction of SHI.

### Descriptive statistics – socio-economic characteristics and current health insurance status

The sample consists of 6,894 individuals of which 2,749 are current or former employees and fall in one of three categories – public sector workers (civil servants or public sector enterprise workers), private sector workers including NGOs, and (former public sector workers) pensioners. The sample consists of 54% public sector workers, 38% private sector workers while the remainder (8%) are pensioners. About half the sample is male and 72% are adults. The average household size is 4.3. Since the sample consists of formal sector workers it is not surprising that about 60% have tertiary education. Education levels are highest amongst public sector workers – 77% have tertiary education as opposed to 73% amongst private sector workers. With regard to monthly income, pensioners have the lowest income while private sector employees record the highest income.

Table 4 provides information on knowledge of insurance and the current health insurance status of sample respondents. While almost all respondents had heard of health insurance, a much smaller fraction – about 40% - was able to provide correct answers to a set of five questions on the functioning of insurance. There is limited variation across public and private sectors. On average, about 52% of households have at least one household member who has health insurance. This figure varies substantially across sectors with 77% of private sector workers reporting that at least one household member has health insurance while the corresponding figures are 21% among pensioners and about 40% amongst public sector workers. For a majority of current employees (about 75%), their employer pays the insurance premium.

**Table 4.**
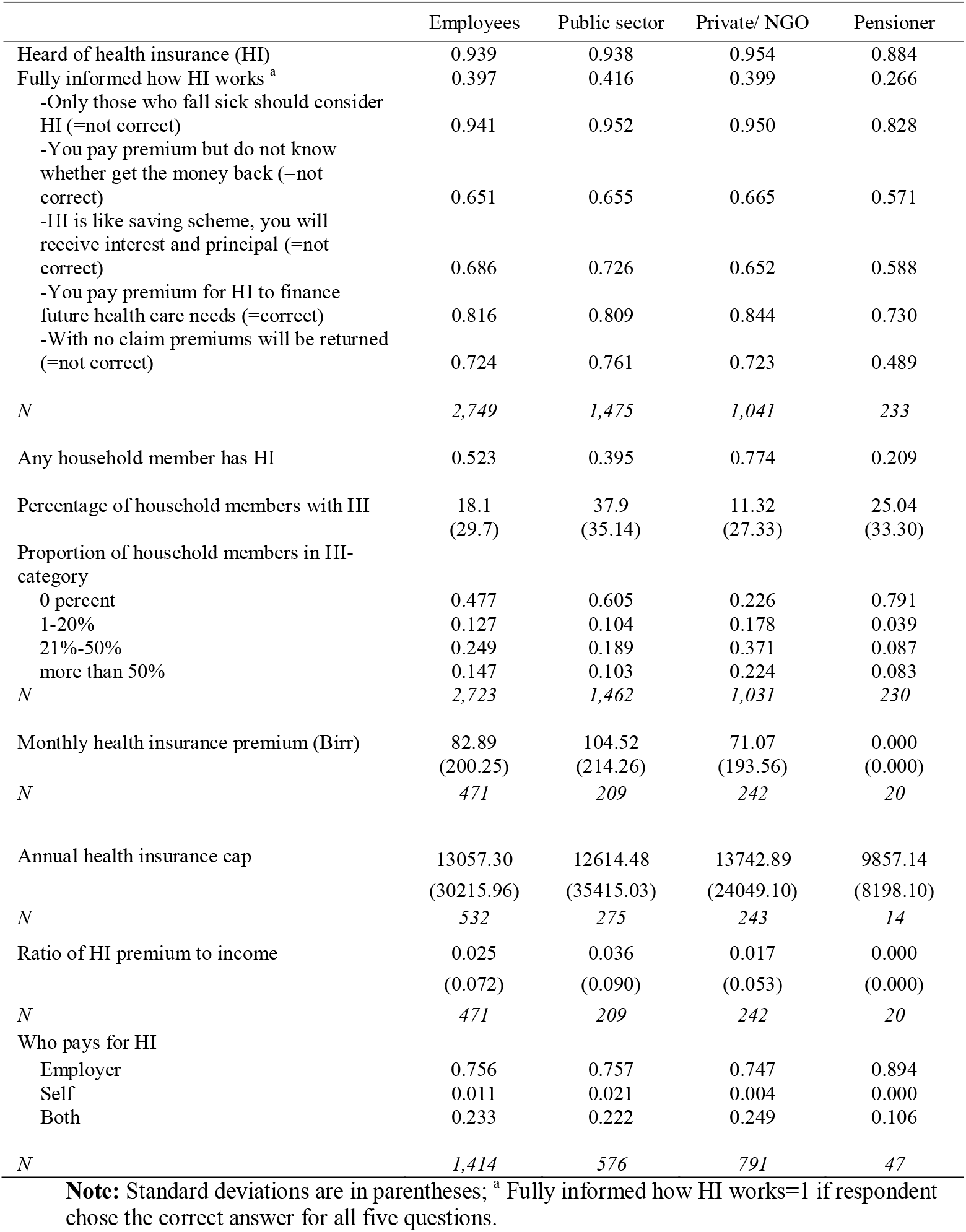
Characteristics related to health insurance

### Seeking outpatient healthcare

Conditional on experiencing an illness in the two months preceding the survey, 85.5% of respondents seek treatment. Amongst those who don’t seek treatment, the majority (more than 50%) expect to recover naturally and hence don’t seek care. Amongst those who do seek care, almost all (94%), opt for formal care. In terms of sector of care, 56% opt for care at a private health facility while the remainder opt for a public facility (see Table 5). The interaction between sector and type of provider is illustrated in Figure 1. A majority of the care is provided by doctors in private facilities (47%) followed by doctors in public facilities (25%), health workers in private facilities (19%) and health workers in the public sector (9%). There are no sharp differences across type of employment. Despite the higher costs of doctor visits to private facilities, where the cost of outpatient care is almost 2.66 times compared to the cost at public facilities, the bulk of formal sector workers opt for private care due to the perceived capability of staff and the availability of drugs while the cheaper cost of care is the main reason for choosing public sector facilities (see Table 6).

**Figure 1:**
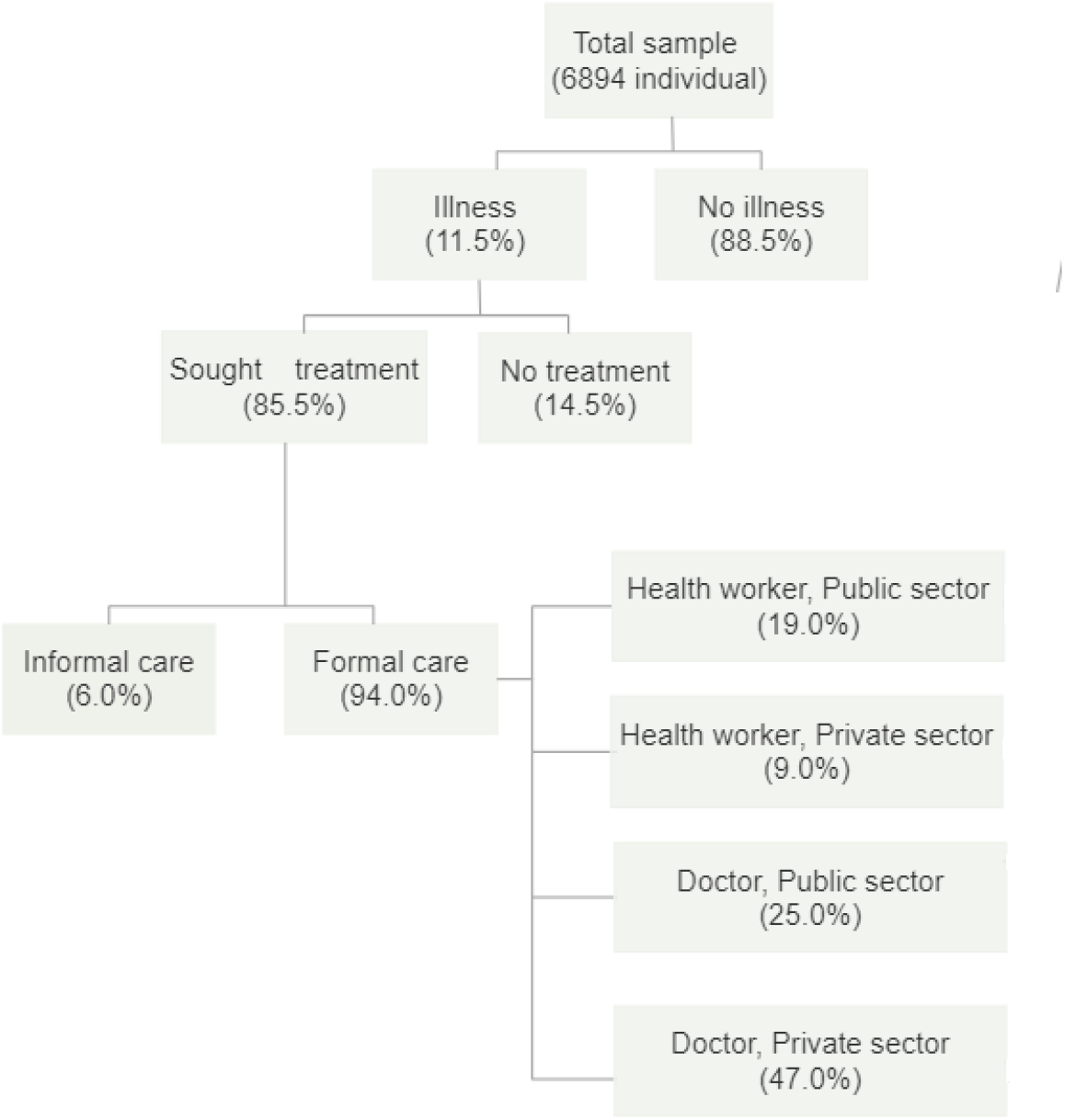
Healthcare seeking choices

**Table 5.** Healthcare seeking

|  | Full sample | Employees | Public sector | Private/ NGO | Pensioner |
| --- | --- | --- | --- | --- | --- |
| Illness or injury in the last two months | 0.115 | 0.145 | 0.139 | 0.139 | 0.206 |
| Seeking any treatment | 0.855 | 0.871 | 0.897 | 0.853 | 0.813 |
| *N* | 779 | 394 | 203 | 143 | 48 |
| Who provided treatment |  |  |  |  |  |
| Informal treatment <sup>a</sup> | 0.061 | 0.057 | 0.090 | 0.008 | 0.054 |
| Self-medication | 0.041 | 0.045 | 0.079 | 0.000 | 0.027 |
| Religious/traditional healer | 0.020 | 0.012 | 0.011 | 0.008 | 0.027 |
| Treatment sector |  |  |  |  |  |
| Public health centre, clinic, or hospital | 0.444 | 0.435 | 0.433 | 0.395 | 0.588 |
| Private health centre clinic, or hospital <sup>b</sup> | 0.556 | 0.565 | 0.567 | 0.605 | 0.412 |
| Treatment type and sector |  |  |  |  |  |
| Health worker- Public sector | 0.190 | 0.169 | 0.186 | 0.134 | 0.212 |
| Health worker- Private sector | 0.089 | 0.110 | 0.083 | 0.160 | 0.061 |
| Doctor-Public sector | 0.250 | 0.263 | 0.244 | 0.261 | 0.364 |
| Doctor- Private sector | 0.470 | 0.458 | 0.487 | 0.445 | 0.364 |
| Treatment cost (Birr) | 745.972<br>(2813.531) | 893.958<br>(3408.822) | 592.294<br>(941.787) | 1366.310<br>(5462.932) | 851.400<br>(2247.338) |
| Delay in seeking treatment (days) | 2.397<br>(4.120) | 2.435<br>(4.603) | 2.650<br>(5.421) | 2.271<br>(3.895) | 2.000<br>(2.000) |
| *N* | 655 | 335 | 177 | 121 | 37 |
| Hospitalization in the last 12 months | 0.018 | 0.025 | 0.024 | 0.030 | 0.013 |
| Inpatient care duration | 9.131<br>(11.977) | 9.043<br>(12.695) | 9.444<br>(15.874) | 8.267<br>(8.103) | 12.000<br>(9.644) |
| *N* | 122 | 69 | 36 | 30 | 3 |
| Inpatient care sector |  |  |  |  |  |
| Public sector | 0.625 | 0.612 | 0.559 | 0.633 | 1.000 |
| Private sector | 0.375 | 0.388 | 0.441 | 0.366 | 0.000 |
| Inpatient care cost (Birr) | 5062.39<br>(6623.97) | 4654.333<br>(5632.223) | 4026.400<br>(5635.470) | 5722.321<br>(5821.649) | 2012.333<br>(438.641) |
| *N* | 118 | 66 | 35 | 28 | 3 |

**Note:** Standard deviations are in parentheses. <sup>a</sup> Informal treatment refers to self-medication, religious and traditional healer. <sup>b</sup>Health worker here refers to health workers, officers, nurses, and midwives. <sup>c</sup> Private sector here includes NGO and missionary facilities.

**Table 6.** Properties of healthcare choices

| Healthcare choice | Two main reasons motivating choice | Satisfied with the treatment | Average cost of treatment (s.d) |
| --- | --- | --- | --- |
| Health worker- Public facilities | Cost of care (41%)<br>Proximity (20.5%) | 81.7% | 199.4<br>(587.9) |
| Health worker- Private facilities | HI covers the cost of treatment (39.6%)<br>Proximity (15.1%) | 92% | 371.0<br>(473.1) |
| Doctor- Public facilities | Cost of care (19.4%)<br>Medicine is available at the location (16.7%) | 83.9% | 476.0<br>(1113.1) |
| Doctor- Private facilities | Capable staff (16.1 %)<br>Medicine is available at the location (15.4%) | 92% | 1265.2<br>(4080.7) |
| Inpatient care- Public facilities | Cost of care (23.1%)<br>Staff are always available (18.5%) | 79.2% | 2360.3<br>(2526.2) |
| Inpatient care- private facilities | Staff are more compassionate (29.3%)<br>Staff are more capable/available (17.1%) | 93.3% | 9358.1<br>(8542.9) |

Table 7 provides a multivariate analysis of the probability of seeking outpatient care (logit model) from a formal provider and subsequently the probability of seeking outpatient care from one of four formal health care providers (multinomial logit). Two points stand out. First, the role of income and second the effect of access to insurance. As compared to the poorest quintile, individuals in the richest income quintile are 5 percentage points (p-value <0.10) more likely to seek formal care. As may be expected, access to insurance translates into a higher probability of using formal care. Residing in a household where a member has health insurance is associated with a 5 percentage (p-value < 0.05) point increase in seeking formal care.

**Table 7.** Probability of seeking outpatient care and choice of outpatient care provider - marginal effects

|  | Seeking<br>treatment | Seeking<br>formal<br>treatment | Choice of formal healthcare provider |  |  |  |
| --- | --- | --- | --- | --- | --- | --- |
|  |  |  | Health worker<br>Public sector | Health worker<br>Private sector | Doctor<br>Public sector | Doctor<br>Private<br>sector |
| Sex (female as reference) | -0.016<br>(0.467) | -0.014<br>(0.485) | 0.009<br>(0.777) | 0.009<br>(0.720) | -0.051<br>(0.163) | 0.033<br>(0.413) |
| Age (under 18 years old as reference) |  |  |  |  |  |  |
| 18-34 years old | -0.045<br>(0.182) | -0.034<br>(0.247) | -0.082<br>(0.083) | 0.072<br>(0.012) | -0.016<br>(0.750) | 0.025<br>(0.642) |
| 35-55 years old | -0.007<br>(0.835) | -0.020<br>(0.404) | -0.102<br>(0.021) | 0.009<br>(0.727) | 0.050<br>(0.308) | 0.042<br>(0.439) |
| 55 years and older | -0.090<br>(0.041) | -0.013<br>(0.670) | -0.145<br>(0.005) | 0.025<br>(0.516) | 0.146<br>(0.047) | -0.025<br>(0.731) |
| Household size (1-person as reference) |  |  |  |  |  |  |
| 2-3 person | 0.047<br>(0.267) | 0.003<br>(0.912) | 0.031<br>(0.539) | 0.084<br>(0.006) | -0.080<br>(0.184) | -0.036<br>(0.598) |
| 4-5 person | 0.061<br>(0.190) | -0.059<br>(0.056) | 0.074<br>(0.186) | 0.026<br>(0.378) | 0.040<br>(0.542) | -0.140<br>(0.046) |
| 6 and more person | 0.091<br>(0.059) | 0.000<br>(0.992) | 0.033<br>(0.612) | 0.072<br>(0.051) | -0.029<br>(0.682) | -0.077<br>(0.344) |
| Education-Household head (no formal edu. as ref.) |  |  |  |  |  |  |
| Primary education | -0.133<br>(0.030) | -0.018<br>(0.756) | 0.005<br>(0.964) | -0.140<br>(0.083) | 0.234<br>(0.006) | -0.099<br>(0.438) |
| Secondary education | -0.124<br>(0.026) | -0.017<br>(0.769) | -0.132<br>(0.242) | -0.103<br>(0.229) | 0.212<br>(0.007) | 0.022<br>(0.859) |
| Tertiary/university education | -0.068<br>(0.161) | 0.004<br>(0.938) | -0.065<br>(0.565) | -0.126<br>(0.107) | 0.109<br>(0.112) | 0.082<br>(0.492) |
| Household income (First quintile(poorest) as ref.) |  |  |  |  |  |  |
| Second quintile | -0.013<br>(0.686) | 0.028<br>(0.370) | -0.039<br>(0.487) | 0.099<br>(0.018) | -0.040<br>(0.487) | -0.019<br>(0.775) |
| Third quintile | 0.016<br>(0.634) | -0.023<br>(0.558) | -0.053<br>(0.364) | -0.009<br>(0.762) | 0.026<br>(0.674) | 0.036<br>(0.603) |
| Fourth quintile | 0.016<br>(0.643) | 0.017<br>(0.639) | -0.045<br>(0.498) | -0.010<br>(0.747) | -0.021<br>(0.746) | 0.076<br>(0.316) |
| Fifth quintile | -0.021<br>(0.624) | 0.052<br>(0.078) | -0.158<br>(0.006) | 0.043<br>(0.251) | -0.134<br>(0.025) | 0.249<br>(0.001) |
| Any household member has HI | 0.020<br>(0.370) | 0.049<br>(0.019) | -0.076<br>(0.019) | 0.022<br>(0.345) | -0.031<br>(0.396) | 0.085<br>(0.039) |
| N | 757 | 645 | 594 |  |  |  |

**Note:** All models include control variables for region (city). P-values are in parentheses.

These patterns are emphasized in the results emerging from the MNL model. The two clearest effects are those related to income and access to health insurance. Individuals belonging to the highest income quintile are far more likely to seek care from private sector providers. For instance, as compared to the poorest quintile, individuals belonging to the richest income quintile are 25 percentage points (p-value <0.01) *more likely* to seek care from doctors in the private sector and 13 percentage points (p-value <0.05) *less likely* to access care from doctors in the public sector. The differences in health-seeking behaviour between the poorest and other income quintiles is not particularly pronounced. Individuals residing in households with access to health insurance are about 9 percentage points (p-value <0.05) more likely to seek care from doctors at private facilities while avoiding the use of public sector health workers (p-value <0.05). We also estimated an alternative specification to examine differences in health seeking behaviour across employees in different sectors (see Table A3). The results indicate that the sector of employment does not influence HSB as opposed to income which remains salient.

### Seeking inpatient healthcare

Approximately 2 percent of the sample has experienced an episode of hospitalization in the 12 months preceding the survey with an average duration of 9 days. In contrast to outpatient care a majority of the sample (62.5%) made use of public facilities (Table 5). On average, the cost of public facilities is about a quarter of that in private facilities and the main attraction of using public facilities is their lower cost. Private facilities are costlier and staff at such facilities are considered more compassionate and capable (Table 6). A logit analysis (Table 8) of the probability of using public care confirms the effect of income on health care choice and shows that households in the higher income quintiles have substantially lower probabilities of using publicly provided care. As compared to the lowest income quintile, respondents in the top three income quintiles are 36 to 56 percentage points (p<0.001) less likely to use public care. The availability of health insurance reduces the probability of using public health care by 26 percentage points (p-value <0.05). The inclusion of sector of work does not alter the role of income in influencing choice of health care provider but does wash out the effect of health insurance suggesting that access to health insurance may be serving as a proxy for working in the private sector.

**Table 8.** Probability of using inpatient care from a public facility - marginal effects

|  | Full sample | Employees |
| --- | --- | --- |
| Sex (female as reference) | 0.120<br>(0.327) | 0.077<br>(0.523) |
| Age (under 18 years old as reference) |  |  |
| 18-34 years old | 0.000<br>(.) | 0.000<br>(.) |
| 35-55 years old | -0.011<br>(0.934) | 0.038<br>(0.806) |
| 55 years and older | -0.011<br>(0.963) | 0.218<br>(0.299) |
| Household size (1-person as reference) |  |  |
| 2-3 person | 0.073<br>(0.663) | 0.061<br>(0.722) |
| 4-5 person | 0.001<br>(0.995) | -0.085<br>(0.612) |
| 6 and more person | 0.007<br>(0.965) | -0.041<br>(0.791) |
| Education <sup>a</sup> (less than secondary education as reference) |  |  |
| Secondary education | 0.031<br>(0.840) | 0.083<br>(0.559) |
| Tertiary/university education | 0.163<br>(0.390) | 0.282<br>(0.137) |
| Income <sup>b</sup> (First quintile(poorest) as reference) |  |  |
| Second quintile | -0.105<br>(0.272) | -0.104<br>(0.292) |
| Third quintile | -0.365<br>(0.003) | -0.487<br>(0.000) |
| Fourth quintile | -0.563<br>(0.000) | -0.550<br>(0.001) |
| Fifth quintile | -0.513<br>(0.000) | -0.489<br>(0.001) |
| Any household member has HI | -0.255<br>(0.034) | -0.174<br>(0.176) |
| Employment Sector (Public sector as reference) |  |  |
| Private sector |  | 0.135<br>(0.259) |
| N | 74 | 63 |

**Note:** All models include control variables for region (city). P-values are in parentheses. <sup>a</sup> Education refers to education level of the head of household for the full sample and to an individual's education in the employees sample. <sup>b</sup> Income refers to household income for the full sample and to an individual's income in the employees sample. As there were insufficient observations in the first category of the 'Education' variable, the first two levels were merged to create a reference category.

### Attitudes towards social health insurance

The available data provides an opportunity to examine, among other issues, willingness to pay for the scheme. Relevant information on the attitude of respondents towards various aspects of SHI are provided in Table 9. While knowledge about the plans for SHI were already widespread (83% of the sample had heard of SHI) in 2016, knowledge about the SHI premium was not widespread (40% knew the premium). Except for pensioners, all others who indicated that they knew the premium had accurate knowledge of the amount of the SHI premium. After being informed about the premium, about 40% considered it fair and a slightly higher proportion (44%) considered the premium to be affordable. The percentages do not differ considerably across type of employment. Notwithstanding the relatively high fairness and affordability rating, only about 24% were willing to pay 3% or more of their monthly income as a premium. The willingness to pay the 3% premium is similar across employment sectors (23.5% versus 28.7% for the public and private sectors, respectively). Only a minority of respondents were unwilling to pay at all (12.8%). While a majority of the sample (67%) supported the introduction of SHI, the average WTP was 1.6% of monthly income ranging from a low of 1.2% for pensioners to a high of 1.7% for private sector workers. A little more than half the sample (53%) is concerned that after paying for SHI they may not get adequate access to health services. The two main concerns are long waiting times and lack of drugs.

**Table 9.** <u>Attitudes towards Social Health Insurance</u>

|  | Employees | Public sector | Private/ NGO | Pensioner |
| --- | --- | --- | --- | --- |
| Heard of Social Health Insurance (SHI) | 0.832 | 0.872 | 0.758 | 0.906 |
| *N* | 2,749 | 1,475 | 1,041 | 233 |
| Aware of SHI covered services/medicines | 0.393 | 0.440 | 0.302 | 0.455 |
| *N* | 2,273 | 1,276 | 786 | 211 |
| Know SHI premium | 0.396 | 0.410 | 0.352 | 0.469 |
| *N* | 2,278 | 1,278 | 789 | 211 |
| Knowledge of monthly amount of the SHI premium in Birr | 132.14 | 139.28 | 157.81 | 19.42 |
|  | (157.14) | (173.26) | (132.81) | (35.44) |
| *N* | 877 | 513 | 270 | 94 |
| Knowledge of SHI premium as a share of income | 0.031 | 0.034 | 0.032 | 0.013 |
|  | (0.036) | (0.039) | (0.031) | (0.027) |
| The SHI premium is fair |  |  |  |  |
| Yes | 0.397 | 0.377 | 0.390 | 0.550 |
| No | 0.399 | 0.441 | 0.345 | 0.341 |
| Don't know | 0.204 | 0.182 | 0.265 | 0.109 |
| *N* | 2,270 | 1,274 | 785 | 211 |
| The SHI premium is affordable |  |  |  |  |
| Disagree | 0.361 | 0.406 | 0.325 | 0.233 |
| Neither agree nor disagree | 0.199 | 0.175 | 0.247 | 0.166 |
| Agree | 0.439 | 0.419 | 0.428 | 0.602 |
| *N* | 2,253 | 1,265 | 777 | 211 |
| SHI WTP as share of income (%) | 1.624 | 1.645 | 1.704 | 1.207 |
|  | (1.307) | (1.196) | (1.516) | (0.996) |
| *N* | 2,163 | 1,210 | 750 | 203 |
| SHI willingness to pay- category |  |  |  |  |
| No willingness to pay | 0.128 | 0.101 | 0.155 | 0.187 |
| Less than 3% of monthly income | 0.629 | 0.664 | 0.559 | 0.680 |
| 3% and more of monthly income | 0.243 | 0.235 | 0.287 | 0.133 |
| Support SHI |  |  |  |  |
| Support | 0.665 | 0.669 | 0.658 | 0.663 |
| Neither support nor oppose | 0.074 | 0.060 | 0.102 | 0.058 |
| Oppose | 0.26 | 0.271 | 0.239 | 0.279 |
| Concerned that after paying for SHI, not receiving adequate healthcare service | 0.529 | 0.564 | 0.512 | 0.381 |
| Concerned about: |  |  |  |  |
| Long waiting time | 0.863 | 0.873 | 0.844 | 0.864 |
| Lack of drugs | 0.837 | 0.859 | 0.799 | 0.827 |
| Lack of adequate diagnosis facilities | 0.724 | 0.726 | 0.715 | 0.753 |
| Quality of staff | 0.660 | 0.642 | 0.684 | 0.704 |
| Availability of staff | 0.517 | 0.528 | 0.484 | 0.593 |
| *N* | 2,270 | 1,274 | 785 | 211 |

**Note:** Standard deviations are in parentheses

Table 10 presents a series of exploratory regressions, *in seriatim*, on the link between various socio-economic traits and the perception that the SHI premium is not fair, that it is not affordable, and that SHI should be opposed. The analysis yields several patterns. First, income and the perception that the SHI premium is unfair is positively correlated. For instance, respondents in the two richest quintiles are 10 percentage points (p-value <0.05) more likely to perceive the premium as unfair as opposed to the lowest income quintile. These two quintiles are not different from the poorest quintile in terms of premium affordability perceptions but still consider the premium unfair. Consistent with the unfair premium perception, the two richest quintiles are 10-13 percentage points (p-value < 0.01) more likely to oppose SHI. In fact, there is greater resistance from all other income groups as compared to the poorest, but the extent of the resistance is higher amongst the two richest quintiles.

**Table 10.** Probability of opposing SHI and that it is unfair, unaffordable, and opposed - (marginal effects)

|  | SHI is<br>not fair | SHI premium is<br>not affordable | Opposing SHI |
| --- | --- | --- | --- |
| Sex (female as reference) | -0.023<br>(0.302) | -0.002<br>(0.926) | -0.009<br>(0.642) |
| Age (under 34 years old as reference) |  |  |  |
| 35-55 years old | 0.046<br>(0.052) | 0.071<br>(0.002) | 0.042<br>(0.047) |
| 55 years and older | 0.024<br>(0.591) | -0.001<br>(0.988) | 0.041<br>(0.324) |
| Household size (1-person as reference) |  |  |  |
| 2-3 person | 0.043<br>(0.198) | 0.005<br>(0.882) | 0.021<br>(0.476) |
| 4-5 person | 0.017<br>(0.610) | -0.015<br>(0.653) | 0.016<br>(0.586) |
| 6 and more person | -0.005<br>(0.905) | -0.006<br>(0.877) | -0.052<br>(0.116) |
| Education (no formal education as reference) |  |  |  |
| Primary education | -0.123<br>(0.393) | 0.077<br>(0.603) | -0.101<br>(0.492) |
| Secondary education | -0.132<br>(0.347) | 0.042<br>(0.766) | -0.142<br>(0.321) |
| Tertiary/university education | -0.110<br>(0.430) | 0.089<br>(0.527) | -0.131<br>(0.359) |
| Income (First quintile(poorest) as reference) |  |  |  |
| Second quintile | 0.062<br>(0.087) | 0.041<br>(0.257) | 0.069<br>(0.026) |
| Third quintile | 0.039<br>(0.284) | 0.026<br>(0.488) | 0.051<br>(0.099) |
| Fourth quintile | 0.108<br>(0.004) | 0.074<br>(0.050) | 0.127<br>(0.000) |
| Fifth quintile | 0.099<br>(0.013) | 0.059<br>(0.134) | 0.101<br>(0.003) |
| Any household member has HI | -0.051<br>(0.031) | -0.016<br>(0.476) | 0.024<br>(0.241) |
| Employment Sector (Public sector as reference) |  |  |  |
| Private/NGO sector | -0.073<br>(0.003) | -0.066<br>(0.006) | -0.030<br>(0.154) |
| Pensioners | -0.092<br>(0.048) | -0.147<br>(0.000) | 0.002<br>(0.967) |
| Aware of SHI covered services/medicines | -0.020<br>(0.348) | -0.059<br>(0.004) | -0.064<br>(0.001) |
| N | 2233 | 2216 | 2189 |

**Note:** All models include control variables for region (city). P-values are in parentheses. As there were insufficient observations in the first category of the 'Age' variable, the first two levels were merged to create a reference category.

## Discussion

This paper was motivated by the challenges underlying the introduction of compulsory SHI in Ethiopia. Key strengths of the paper include the availability of data representative of formal sector workers residing in four of the country’s main cities, coverage of both private and public sector employees and information which allowed us to examine HSB, attitudes toward SHI, and willingness to pay for SHI. Covering these issues yields a more complete picture of the issues which may be encountered by the government as it sets out to implement SHI.

Regarding the use of outpatient care, conditional on falling ill, 85.5% of the respondents sought care within a couple of days (2.4 days) of falling ill. Almost all respondents (94%) sought formal care with a majority of the care (55.6%) being provided by private health clinics or hospitals. Income and access to health insurance were the two most important factors determining choice of health care provider with richer households and those with health insurance more likely to opt for care from doctors in private facilities. The private sector was preferred due to the perception of more capable staff and availability of drugs. This is consistent with studies on patient satisfaction which demonstrate that patients perceive that they are more likely to receive patient-centred and higher-quality health services in the private sector [14, 15, 18, 27-28]. In the case of inpatient care, the picture was the opposite. A majority of health care visits were to public sector hospitals (62.5%). Given the cost of accessing inpatient care at private facilities (4 times that of public care), the greater reliance on the public sector for inpatient care is not surprising. The results show that it is not the employment sector that matters but income and arguably the position of respondents within their sector of employment that determines their healthcare choices. Despite the lower costs, the perception of lower quality public sector health care drives higher-income households to the private sector while lower-income households have little choice but to visit public health facilities.

The low level of health insurance coverage combined with the relatively expensive care offered by doctors at private facilities suggests that a substantial proportion of health care costs must be financed through out-of-pocket (OOP) expenditure. Despite this burden, the willingness to pay the proposed SHI premium of 3% of monthly income is low. About 24% of formal sector respondents were willing to pay this premium and the average WTP was 1.6% of monthly income. These figures are similar to those reported in the bulk [10-17] of the existing literature. A quarter of respondents (26%) were opposed to the introduction of SHI. The analysis showed that higher income respondents were more likely to oppose the introduction of SHI.

This pattern of results suggests that the low WTP and opposition to SHI is driven not only by affordability concerns, but other concerns as well. There are two main concerns. First, concerns about low-quality services at public facilities. While the use of SHI is not expected to be restricted only to public facilities, it is most likely that EHIA will initially contract mainly public facilities which reduces the attractiveness of SHI. Second, is the fear of double payment. Since SHI is expected to be mandatory it raises the possibility that formal sector workers have to pay SHI premiums while still paying out-of-pocket for the use of health care services at private facilities. The interaction between the low-quality of health care at public facilities and the additional cost of accessing care translates into an unwillingness to pay the 3% premium even amongst those respondents who find it affordable. Conversely, the availability of higher quality care (more patient-centred, availability of drugs and equipment) is likely to increase the willingness to pay the SHI premium [29]. Although in the context of the CBHI, the importance of investing in quality of care – specifically, the availability of drugs and equipment – is not lost on the government [30].

## Conclusion

One of the key reasons for the lack of implementation of the SHI despite the announcement of the policy in the government’s official gazette in 2010 has been the affordability of the proposed SHI premium. The analysis of healthcare usage in this paper suggests that in addition to affordability, the interaction between the quality of health care on offer at public facilities and the willingness to pay for the available quality even amongst those for whom the premium is affordable, reduces support for SHI. Concerns about the double burden of expenditure (payment of SHI premium and no reductions in OOP expenditure) and access to healthcare coverage only at public facilities makes the SHI unattractive even if affordability is not an issue.

What are the policy implications of these findings? First, given the predominant use of private facilities, especially for outpatient care, it is imperative that the EHIA signs contracts with private providers or at the very least articulates the intention to sign contracts with private providers even if it does not happen at program inception. Even if the proposed SHI covers a part of the cost of accessing private care it should be an attractive proposition as compared to the current situation where health insurance coverage is limited, and a substantial proportion of expenditure is financed out-of-pocket. Second and perhaps most importantly, prior to SHI launch, and as was the case prior to the introduction of the voluntary CBHI schemes, concerted efforts to enhance the quality of care – both, in terms of perception (patient-centred care) and in terms of addressing actual bottlenecks (waiting time to see a professional, availability of drugs and equipment) – are needed to enhance support for SHI. The strong link between income and use of private health care facilities underlines the idea that richer respondents are more likely to oppose the introduction of SHI. A combination of ensuring access to private healthcare facilities combined with investments in the quality of health care offered at public facilities is most likely to reduce opposition to the introduction of SHI.

## Data Availability

All data produced in the present study are available upon reasonable request to the authors

## Contributor statement

**ZZ** Data curation, Data analysis, Methodology, Writing – Original draft**; AM** Conceptualization, Survey design, Sampling approach, Data collection, Data curation, Writing – Reviewing and editing; **DS** Survey design, Sampling approach, Data collection, Data curation, Writing – Reviewing and editing; **ZS** Conceptualization, Funding acquisition, Writing – Review and Editing **GA** Conceptualization, Survey design, Sampling approach, Data collection, Project administration, Writing – Reviewing and editing; **AB** Conceptualization, Survey design, Sampling approach, Funding acquisition, Writing – Original Draft

All authors confirm full access to all data used in the study and accept responsibility for the journal submission.

## Data sharing

The data set used for this study will be uploaded in a public repository upon acceptance.

## Appendix Tables

**Table A1.** Distribution of civil servants in Addis Ababa

|  | **Names of organization** | **Total employees** | **Sample size** |
| --- | --- | --- | --- |
| Addis Ababa City Administration | Addis Ababa Road Authority | 2543 | 36 |
|  | Disaster Preparedness and Control Authority | 813 | 12 |
|  | Bureau of Culture and Tourism | 518 | 7 |
|  | Bureau of Women and Children | 470 | 7 |
|  | Mass Media Agency | 374 | 5 |
| Oromia Regional Administration (based in Addis Ababa) | Auditor General | 298 | 4 |
|  | Bureau of Agriculture | 332 | 5 |
|  | Water, Mineral and Energy Bureau | 288 | 4 |
|  | Bureau of Labour & Social Affairs | 300 | 4 |
|  | Bureau of Health | 266 | 4 |
| Federal | Ministry of Agriculture | 1401 | 20 |
|  | Ministry of Health | 1084 | 15 |
|  | Ministry of Water and Energy | 990 | 14 |
|  | Ministry of Education | 830 | 12 |
|  | Ministry of Urban development and Construction | 563 | 8 |
|  | Ministry of Finance and Economic Development | 804 | 11 |
|  | Ministry of Foreign Affairs | 866 | 12 |
|  | EARI (EARO) | 3645 | 52 |
|  | Ethiopian Road Authority | 1396 | 20 |
|  | Governmental House Agency | 1685 | 24 |
|  | Paulos hospital | 1479 | 21 |
|  | Addis Ababa University | 3324 | 47 |
|  | Ethiopian Radio and Television Agency | 2011 | 28 |
| **Total** |  | **26280** | **372** |

**Table A2.** Addis Ababa pension paying centers

| **Sub city** | **Selected payment center** | **Sample Size** |
| --- | --- | --- |
| Yeka | Woreda 8 | 31 |
|  | Tesfa Birhan secondary school |  |
| Gulele | Post office around Paulos hospital close to CBE branch | 31 |
|  | Addis Credit and saving in the premises of Woreda 9 office Rufael |  |
| Bole | Bole Mikael post office (office in a condominium a little down from Bole ring road square) | 31 |
|  | Gerji post office sunshine condominium close to Roba bakery |  |
| Akaki | Saris - Addis Sefer in the premises of Woreda 8 | 31 |
| Nefas Silk Lafto | Post office in Gotera condominium | 31 |
| Total |  | 155 |

**Table A3.** Determinants of health seeking and the choice of provider among formal sector employees (marginal effects)

|  | Seeking treatment | Seeking formal treatment | Choice of formal healthcare provider |  |  |  |
| --- | --- | --- | --- | --- | --- | --- |
|  |  |  | Health worker Public sector | Health worker Private sector | Doctor Public sector | Doctor Private sector |
| Sex (female as reference) | -0.047<br>(0.116) | -0.043<br>(0.235) | 0.006<br>(0.884) | 0.012<br>(0.759) | 0.029<br>(0.588) | -0.048<br>(0.388) |
| Age (18-34 years old as reference) |  |  |  |  |  |  |
| 35-55 years old | 0.018<br>(0.627) | 0.030<br>(0.486) | 0.032<br>(0.504) | -0.064<br>(0.143) | 0.062<br>(0.271) | -0.030<br>(0.618) |
| 55 years and older | -0.063<br>(0.449) | 0.030<br>(0.655) | 0.026<br>(0.802) | -0.116<br>(0.033) | 0.056<br>(0.584) | 0.034<br>(0.781) |
| Household size (1-person as ref.) |  |  |  |  |  |  |
| 2-3 person | 0.093<br>(0.053) | 0.042<br>(0.241) | 0.017<br>(0.783) | 0.133<br>(0.003) | -0.067<br>(0.346) | -0.083<br>(0.302) |
| 4-5 person | 0.123<br>(0.022) | -0.048<br>(0.375) | 0.030<br>(0.652) | 0.036<br>(0.333) | 0.070<br>(0.365) | -0.135<br>(0.095) |
| 6 and more person | 0.161<br>(0.001) | 0.001<br>(0.991) | -0.030<br>(0.699) | 0.082<br>(0.135) | 0.018<br>(0.833) | -0.070<br>(0.480) |
| Education (less than secondary education as ref.) |  |  |  |  |  |  |
| Secondary education | -0.092<br>(0.105) | -0.013<br>(0.770) | 0.037<br>(0.666) | -0.071<br>(0.473) | -0.020<br>(0.838) | 0.055<br>(0.595) |
| Tertiary/university education | 0.022<br>(0.639) | -0.018<br>(0.644) | -0.063<br>(0.426) | -0.064<br>(0.570) | -0.106<br>(0.256) | 0.232<br>(0.024) |
| Income (First quintile as reference) |  |  |  |  |  |  |
| Second quintile | 0.029<br>(0.542) | 0.037<br>(0.584) | 0.025<br>(0.750) | 0.043<br>(0.547) | -0.136<br>(0.117) | 0.068<br>(0.494) |
| Third quintile | 0.016<br>(0.748) | 0.028<br>(0.646) | -0.045<br>(0.536) | -0.006<br>(0.923) | 0.116<br>(0.253) | -0.065<br>(0.520) |
| Fourth quintile | 0.014<br>(0.768) | 0.066<br>(0.256) | -0.021<br>(0.776) | 0.023<br>(0.727) | -0.096<br>(0.296) | 0.095<br>(0.356) |
| Fifth quintile | 0.040<br>(0.469) | 0.056<br>(0.322) | -0.052<br>(0.487) | -0.019<br>(0.795) | -0.212<br>(0.021) | 0.282<br>(0.009) |
| Any household member has HI | -0.018<br>(0.564) | 0.002<br>(0.961) | -0.037<br>(0.422) | 0.065<br>(0.146) | -0.039<br>(0.488) | 0.012<br>(0.855) |
| Employment Sector (Public sector as ref.) |  |  |  |  |  |  |
| Private/NGO sector | 0.000<br>(0.998) | 0.104<br>(0.001) | -0.037<br>(0.445) | 0.030<br>(0.500) | 0.081<br>(0.167) | -0.074<br>(0.213) |
| Pensioners | -0.004<br>(0.947) | 0.072<br>(0.098) | -0.044<br>(0.589) | -0.000<br>(0.998) | -0.024<br>(0.772) | 0.067<br>(0.579) |
| N | 389 | 269 | 308 |  |  |  |

**Note:** All models include control variables for region (city). P-values are in parentheses. As there were insufficient observations in the first category of the 'Education' variable, the first two levels were merged to create a reference category.

## Footnotes

1 Based on previous studies that have examined the effect of Ethiopia’s community-based health insurance on utilization of inpatient care, utilization of outpatient care and OOP expenditure the effect sizes were set at 2.3 percentage points, 5 percentage points and 2.2 percentage points, respectively.

## References

1. FMoH, 2001. Health and Health Related Indicators 1994 E.C (2001/02). https://e-library.moh.gov.et/library/wp-content/uploads/2021/07/Health-and-Health-Related-Indicators-1994-E.C.pdf, Accessed date: 6 August 2021.

2. FMoH, 2021. Health and Health Related Indicators 2013 EFY (2020/21). https://www.moh.gov.et/site/sites/default/files/2021-10/Health%20and%20health%20related%20indicators%20_2013%20EFY.pdf, Accessed date: 5 May 2023.

3. FMoH, 2022. Ethiopia national health accounts report 2019/20. https://apps.who.int/nha/database/DocumentationCentre/GetFile/60033121/en, Accessed date: 5 May 2023.

4. FMoH, 2000. Health and Health Related Indicators 1992 E.C (2001/02). https://e-library.moh.gov.et/library/wp-content/uploads/2021/07/Health-and-Health-Related-Indicators-1992-E.C.pdf, Accessed date: 7 August 2021.

5. FMoH, 2021. Ethiopian Health Service Coverage -Fact Sheet 2020. https://www.moh.gov.et/site/fact-sheets, Accessed date: 5 May 2023.

6. EHIA, 2021. CBHI Members Trend. Presentation.

7. FMoH, 2015. Health Sector Transformation Plan. https://extranet.who.int/nutrition/gina/sites/default/filesstore/ETH%202016%20Health%20Sector%20Transformation%20Plan.pdf, Accessed date: 13 December 2022.

8. FDRE, 2010. Social Health Insurance proclamation No.690/2010. https://www.ilo.org/dyn/travail/docs/269/Social%20Health%20Insurance%20Proclamation%202010.pdf, Accessed date: 12 December, 2022.

9. Obse, A., Hailemariam, D., Normand, C. 2015. Knowledge of and preferences for health insurance among formal sector employees in Addis Ababa: a qualitative study. BMC Health Serv. Res. 15, 318.

10. Kokebie, M.A., Abdo, Z.A., Mohamed, S., Leulseged, B. 2022. Willingness to pay for social health insurance and its associated factors among public servants in Addis Ababa, Ethiopia: a cross-sectional study. BMC Health Serv. Res. 22(1):909.

11. Lasebew, Y., Yeshwondm, M., Abdelmenan, S. 2017. Willingness to Pay for the Newly Proposed Social Health Insurance among Health Workers at St. Paul’s Hospital Millennium Medical College, Addis Ababa, Ethiopia. International Journal of Health Economics and Policy 2 (4):159–166.

12. Mekonne, A., Seifu, B., Hailu, C., Atomsa, A. 2020. Willingness to Pay for Social Health Insurance and Associated Factors among Health Care Providers in Addis Ababa, Ethiopia. Biomed Res Int. 2020: 8412957

13. Obse A, Ryan M, Heidenreich S, Normand C, Hailemariam D. 2016. Eliciting preferences for social health insurance in Ethiopia: a discrete choice experiment. Health Policy Plan. 31(10):1423–1432.

14. Yeshiwas, S., Kiflie, M., Zeleke, A.A., Kebede, M. 2018. Civil servants’ demand for social health insurance in Northwest Ethiopia. Arch Public Health 76, 48.

15. Zemene, A., Kebede, A., Atnafu, A., Gebremedhin, T. 2020. Acceptance of the proposed social health insurance among government-owned company employees in Northwest Ethiopia: implications for starting social health insurance implementation. Arch Public Health 78, 104.

16. Amilaku, E., Fentaye F., Mekonen, A., Bayked, E. 2022. Willingness to pay for social health insurance among public civil servants: A cross-sectional study in Dessie City Administration, North-East Ethiopia. Front Public Health. 10:920502.

17. Mekonnen, F., Agumas, Y., Mulu, Y., Fentaw, G., Wassie, F. 2021. Willingness to Pay for Social Health Insurance and Its Predictors among Government Employees in Mujja Town, Ethiopia. Scientific World Journal. 2021:3149289.

18. Gidey, M., Gebretekle, G., Hogan, M., Fenta, T. 2019. Willingness to pay for social health insurance and its determinants among public servants in Mekelle City, Northern Ethiopia: a mixed methods study. Cost Eff Resour Alloc. 17:2.

19. Agago, T., Woldie, M., Ololo, S. 2014. Willingness to join and pay for the newly proposed social health insurance among teachers in Wolaita Sodo Town, South Ethiopia. Ethiop J Health Sci. 24(3):195–202.

20. Setegn A, Andargie G, Amare G, Debie A. 2021. Willingness to Pay for Social Health Insurance Among Teachers at Governmental Schools in Gondar Town, Northwest Ethiopia. Risk Manag Healthc Policy 14:861–868.

21. Lavers, T. 2019. Towards Universal Health Coverage in Ethiopia’s ’developmental state’? The political drivers of health insurance. Soc Sci Med. 228:60–67.

22. Begashaw B, Tessema F, Gesesew HA. 2016. Health Care Seeking Behavior in Southwest Ethiopia. PLoS ONE 11(9): e0161014

23. Deressa W, Seme A, Asefa A, Teshome G, Enqusellassie F. 2014. Utilization of PMTCT services and associated factors among pregnant women attending antenatal clinics in Addis Ababa, Ethiopia. BMC Pregnancy Childbirth. 4:328.

24. Bantie GM, Meseret Z, Bedimo M, Bitew A. 2019. The prevalence and root causes of delay in seeking healthcare among mothers of under five children with pneumonia in hospitals of Bahir Dar city, North West Ethiopia. BMC Pediatr. 19(1):482.

25. Gelaw YA, Biks GA, Alene KA. 2014. Effect of residence on mothers’ health care seeking behavior for common childhood illness in Northwest Ethiopia: A community based comparative cross--sectional study. BMC Res Notes 7:705.

26. Tekalign T, Guta MT, Awoke N, Asres AW, Obsa MS. 2022. Mothers’ Care-Seeking Behavior for Common Childhood Illnesses and Its Predictors in Ethiopia: Meta-Analysis. Int J Pediatr. 2022:2221618.

27. Biresaw H, Mulugeta H, Endalamaw A, Yesuf NN, Alemu Y. 2021. Patient satisfaction towards health care services provided in Ethiopian Health Institutions: A systematic review and meta-analysis. Health Services Insights 14:1–12.

28. Birhanu F, Yitbarek K, Addis A, Alemayehu D, Shifera N. 2021. Patient-centered care and associated factors at public and private hospitals of Addis Ababa: Patients’ perspective. Patient Related Outcome Measures. 12:107–16.

29. Alemtsehay T, Yitayal M, Kebede A. 2020 Acceptance for Social Health Insurance among Health Professionals in Government Hospitals, Mekelle City, North Ethiopia, Advances in Public Health, https://doi.org/10.1155/2020/6458425.

30. Ethiopian Health Insurance Agency. 2020. CBHI Members’ Registration and Contribution 2011_j2020. CBHI Trend Bulletin.

